# SPATIAL ANALYSIS OF MORTALITY DUE TO CONGENITAL SYPHILIS IN BRAZIL FROM 2008 TO 2022

**DOI:** 10.1101/2025.06.01.25328756

**Authors:** Yago Tavares Pinheiro, Richardson Augusto Rosendo da Silva, Ketyllem Tayanne da Silva Costa, Angelo Giuseppe Roncalli da Costa Oliveira, Janmilli Dantas da Costa, Cristiane da Silva Ramos Marinho, Jurandir Alves de Freitas Filho, Luennia Kerlly Alves Rocha, Victória Sampaio Moreira, Ruan Carlos de Queiroz Monteiro, José Rebberty Rodrigo Holanda

**Affiliations:** Department of Public Health, Federal University of Rio Grande do Norte, Natal, Brazil; Faculty of Health Science of Trairí, Federal University of Rio Grande do Norte, Natal, Brazil; Centro Universitário Santa Maria, Cajazeiras, Paraíba, Brazil; Escola Multicampi de Ciências Médicas do Rio Grande do Norte, Federal University of Rio Grande do Norte, Natal, Brazil

**Keywords:** Sexually transmitted infections, Mortality, Spatial analysis, Epidemiology

## Abstract

The objective of this study is to analyze spatial distribution of mortality due to congenital syphilis in Brazil from 2008 to 2022. This is an ecological study that considered congenital syphilis deaths reported in all Brazilian municipalities, from 2008 to 2022, available in the Brazilian government’s information systems. We built a thematic map to describe the distribution of congenital syphilis mortality in the country and, subsequently, applied the Local Index Spatial Analysis to identify possible spatial clusters. Finally, we used the Ordinary Least Squares and Geographically Weighted Regression regression models to identify mortality predictors in the territory. The mortality rate from congenital syphilis was 0.64 deaths per 1,000 live births. The distribution of deaths occurred heterogeneously, with the highest rates in the states of Pará, Acre, Rondônia, Rio de Janeiro and part of Amazonas. We identified statistically significant spatial clusters across the country, with the formation of clusters with a high-high pattern in Pará, Rio de Janeiro, and Mato Grosso (p<0.05). We observed that the Gini index (p=0.008; 95% Confidence Interval: 0.02 – 0.11), the number of nurses in primary care (p=0.027; 95% Confidence Interval: 0.0005 – 0.00003) and the proportion of non-treponemal tests by pregnant women (p=0.016; 95% Confidence Interval: 0.005 – 0.001) are variables that influence the occurrence of deaths. Congenital syphilis deaths in Brazil occur heterogeneously, with different rates between regions, which are geographically influenced by social and healthcare characteristics of the location.

## Introduction

The number of deaths from congenital syphilis (CS) in Brazil has increased significantly in recent years [1]. Between 2011 and 2021, there was a 39.9% rise in deaths from the disease nationwide. Moreover, during the same period, the infant mortality rate due to CS increased by 84.6%, rising from 3.8 to 7.0 deaths per 100,000 live births [2]. Some studies have estimated infant and neonatal mortality due to CS in Brazil, reporting an increase in the mortality rate from 2.74 to 7.84 deaths per 100,000 live births between 2010 and 2017 [3,4].

In addition, it was observed that the risk of death was higher among children born to untreated mothers or mothers with low educational attainment, in cases with treponemal titers greater than 1:64, and when clinical signs and symptoms were present at birth [1,3,4]. However, despite the currently available literature, there remains a lack of data on the geographic distribution of CS-related deaths in Brazil, as well as on the factors influencing their spatial dynamics. This gap hinders the identification of areas most vulnerable to such outcomes and limits the ability to target control strategies to regions in greatest need.

In this context, analyzing the geographic distribution of CS-related deaths and identifying their influencing factors—based on the most recent data provided by the Brazilian government over a broad historical period (2008 to 2022)—may yield valuable information to support the identification of areas with a higher propensity for such deaths. This, in turn, can facilitate the development and implementation of more effective preventive interventions tailored to the specific characteristics of each region. Furthermore, the results of such analyses may serve as a reference for other countries with similar conditions to define and implement their own strategies for controlling CS-related deaths.

Given this scenario, an in-depth analysis of the epidemiological situation is necessary to generate evidence that can support the design and implementation of effective strategies for reducing CS-related mortality in Brazil. Therefore, the objective of this study was to analyze the spatial distribution of congenital syphilis mortality in Brazil from 2008 to 2022.

## Methods

This is an ecological study, developed using secondary data from all municipalities in Brazil. The country comprises 5,570 municipalities, distributed across 27 federative units (26 states and one Federal District). Geographically, it is divided into five macro-regions: North, Northeast, Southeast, South, and Central-West [5]. The study population consisted of newborns (aged 0 to 27 days) diagnosed with congenital syphilis in Brazil.

All data were collected in July 2023 from databases linked to the Brazilian Ministry of Health, the Department of Informatics of the Unified Health System (DATASUS), the United Nations Development Programme (UNDP), and the Brazilian Institute of Geography and Statistics (IBGE). The data covered a fifteen-year period, from 2008 to 2022.

For this study, the following data were collected: the number of deaths due to congenital syphilis among newborns aged 0 to 27 days by municipality of residence at the time of notification, the number of live births, the percentage of non-treponemal tests performed on pregnant women, and the proportion of primary care nurses per inhabitant. The definitions of the variables are described in the variable matrix (Table 1).

**Table 1.**
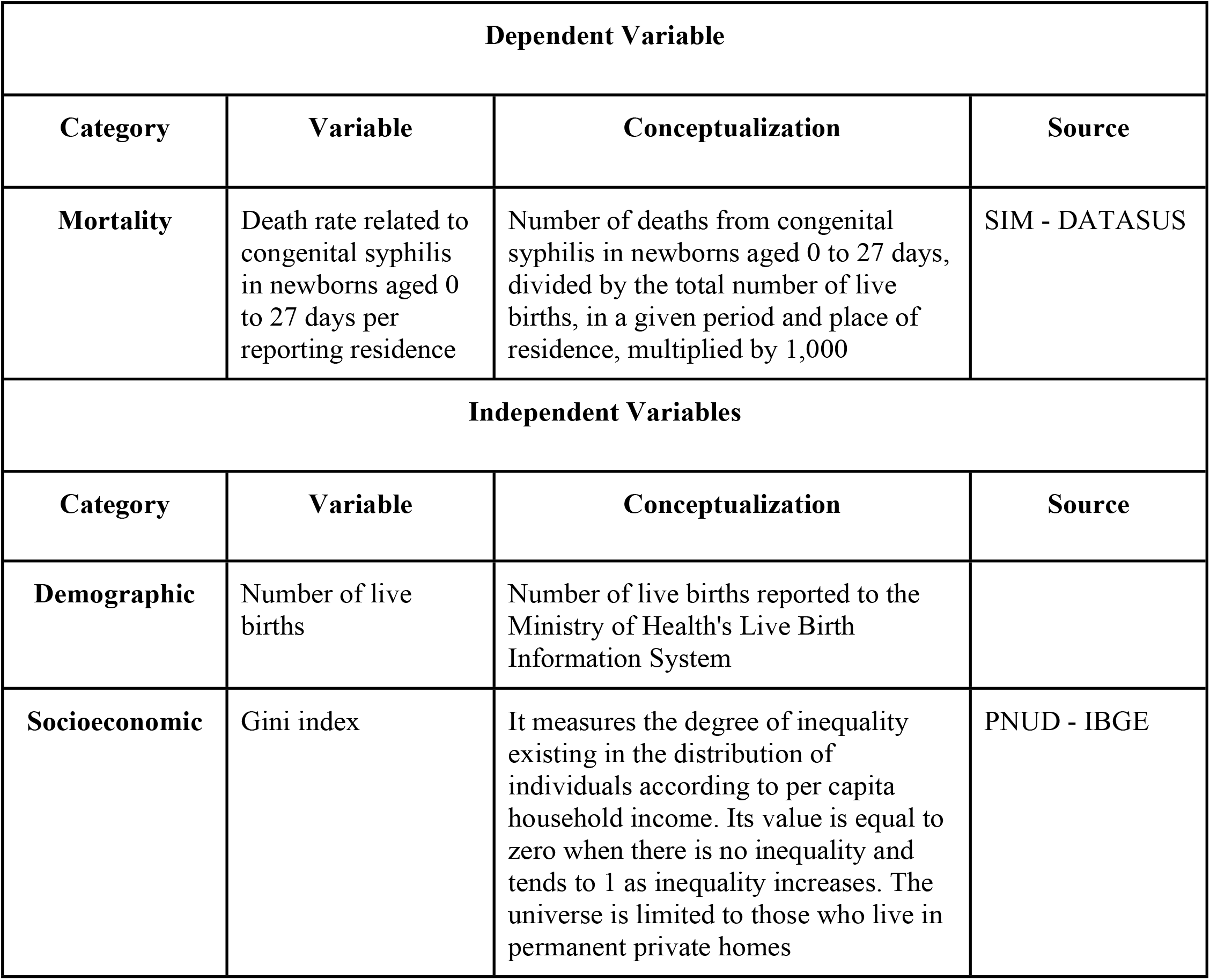
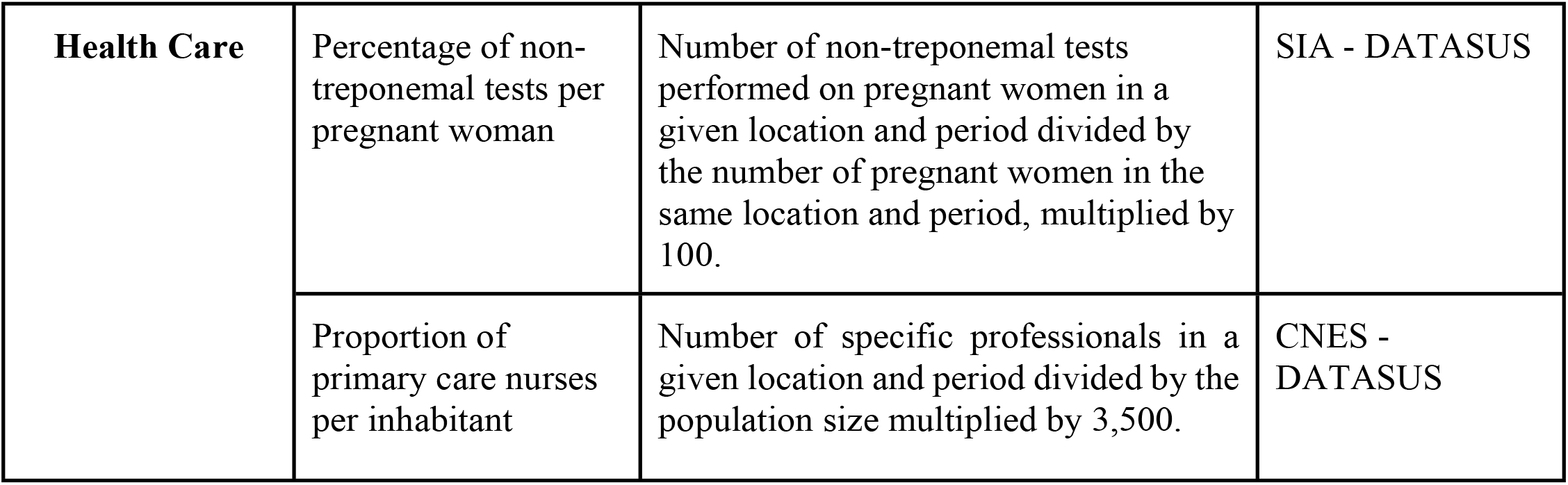
Characterization of the dependent and independent variables used in the statistical analysis of the study. Brazil, 2025.

| Dependent Variable |  |  |  |
| --- | --- | --- | --- |
| Category | Variable | Conceptualization | Source |
| Mortality | Death rate related to congenital syphilis in newborns aged 0 to 27 days per reporting residence | Number of deaths from congenital syphilis in newborns aged 0 to 27 days, divided by the total number of live births, in a given period and place of residence, multiplied by 1,000 | SIM - DATASUS |
| Independent Variables |  |  |  |
| Category | Variable | Conceptualization | Source |
| Demographic | Number of live births | Number of live births reported to the Ministry of Health's Live Birth Information System |  |
| Socioeconomic | Gini index | It measures the degree of inequality existing in the distribution of individuals according to per capita household income. Its value is equal to zero when there is no inequality and tends to 1 as inequality increases. The universe is limited to those who live in permanent private homes | PNUD - IBGE |
| **Health Care** | Percentage of non-treponemal tests per pregnant woman | Number of non-treponemal tests performed on pregnant women in a given location and period divided by the number of pregnant women in the same location and period, multiplied by 100. | SIA - DATASUS |
|  | Proportion of primary care nurses per inhabitant | Number of specific professionals in a given location and period divided by the population size multiplied by 3,500. | CNES - DATASUS |

The outcome variable was the average mortality rate due to congenital syphilis (aged 0 to 27 days), standardized by the population of live births in the year 2015, which represents the median year of the study period. The remaining variables were considered as potential predictors of the outcome variable. The congenital syphilis mortality rate was calculated for each municipality in Brazil using the following equation:

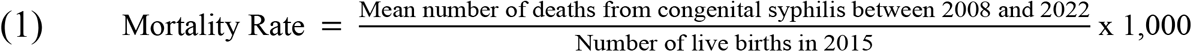

For the spatial analysis, a thematic map of congenital syphilis mortality in Brazilian municipalities was initially created. Subsequently, crude mortality rates were smoothed using the local empirical Bayesian method to reduce instabilities caused by municipalities with no reported cases or rates that were markedly different from those of neighboring areas. The application of this method is necessary because it produces rates that are more representative of reality, as it accounts not only for the value of a given municipality but also weights it in relation to its neighbors, using a spatial proximity matrix. This matrix was constructed based on a contiguity criterion, assigning a value of 1 to municipalities that share borders and 0 to those that do not [6].

Following the analysis of spatial distribution of deaths, spatial clusters were identified using Moran’s spatial autocorrelation function. The Local Moran’s Index (Local Indicators of Spatial Association – LISA) was applied to detect spatial clusters and quantify the degree of spatial association in each municipality of the sample, with statistical significance set at p<0.05 [7].

The results of the Local Moran’s Index were visualized through a Moran Map. This map graphically displays the degree of similarity between neighboring areas and is represented by four quadrants: in quadrant one (Q1), municipalities with high rates adjacent to other high-rate municipalities are shown (High-High spatial pattern); in quadrant two (Q2), municipalities with low rates surrounded by other low-rate municipalities are displayed (Low-Low spatial pattern); quadrant three (Q3) includes municipalities with high rates surrounded by low-rate neighbors (High-Low spatial pattern); and quadrant four (Q4) represents municipalities with low rates surrounded by high-rate neighbors (Low-High spatial pattern) [7]. In addition to identifying spatial clustering quadrants, the significance of these clusters can also be determined.

Following this, a purely spatial scan analysis was performed using the Scan statistical technique. This method allows for the identification of high-risk areas for both outcomes in comparison to Brazil as a whole. Purely spatial clusters were identified using the discrete Poisson model with the following parameters: no geographic overlap of clusters, a maximum cluster size of 50.0% of the exposed population, circular cluster shapes, and 999 replications. Relative risk was calculated for each municipality, where values >1 indicated a higher risk of congenital syphilis mortality compared to the national average [8].

Finally, an analysis was conducted to identify predictors of congenital syphilis mortality. For this analysis, an Ordinary Least Squares (OLS) regression model was used to assess the relationship between variables through a stepwise method, with a removal threshold of p>0.10. The results were compared with those of a geographically weighted regression (GWR) model, which tests the hypothesis of spatially varying relationships among variables. The model with the best fit was selected based on the adjusted R^2^ and the Akaike Information Criterion (AIC) [9].

Data tabulation and proportion calculations were performed in Microsoft Excel 2016. The local empirical Bayesian rate and spatial autocorrelation tests were carried out using GeoDa 1.14 software. The purely spatial scan analysis was conducted in SaTScan 9.7, and the geographically weighted regression analysis was performed in GWR 4. All maps were created using QGIS 3.16 software.

The data used in this study are publicly available, and therefore, ethical approval was not required, in accordance with Resolution No. 466/12 of the Brazilian National Health Council.

## Results

Between 2008 and 2022, a total of 214,203 cases of congenital syphilis were reported in Brazil, of which 1,927 resulted in death due to complications occurring within the first 0 to 27 days of life, among a population of 2,996,967 live births. Thus, the national mean mortality rate was 0.64 deaths per 1,000 live births. The distribution of mortality was heterogeneous across the country; in most municipalities, the rate ranged between 0.0 and 0.1 per 1,000 live births. However, in some municipalities, it exceeded 1 per 1,000 live births (Figure 1A). The application of the local empirical Bayesian method enabled a clearer visualization of the spatial distribution of this indicator, especially in the states of Pará, Rio de Janeiro, parts of Amazonas, Acre, and Rondônia (Figure 1B).

**Figure 1.**
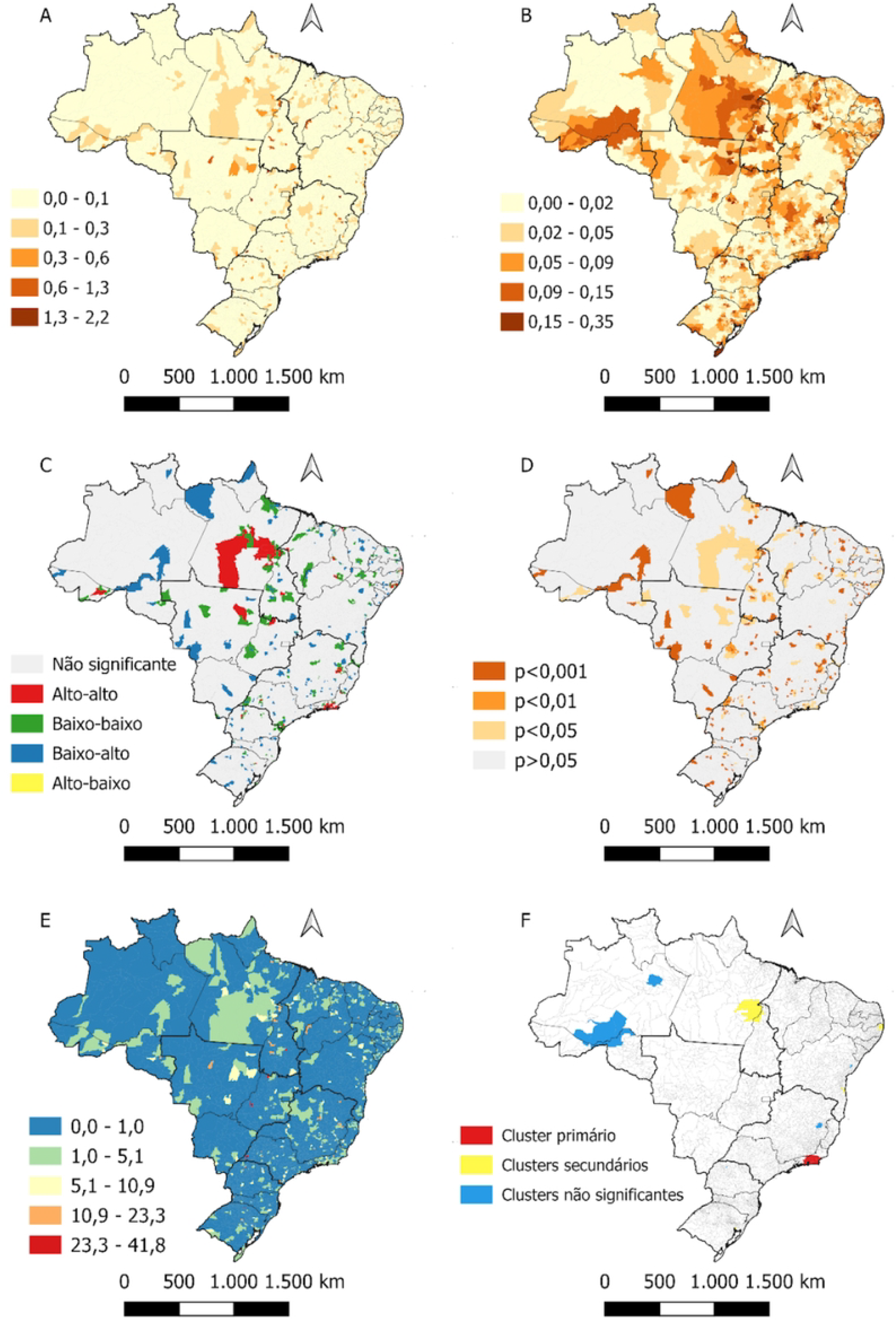
Spatial pattern of deaths from congenital syphilis in Brazil from 2008 to 2022.

The Local Moran’s Index allowed the identification of spatial clusters throughout the Brazilian territory. High-high clusters were observed in Pará, Rio de Janeiro, Mato Grosso, and, to a lesser extent, in other states (Figure 1C). All clusters showed statistical significance (p<0.05) (Figure 1D).

The application of spatial scan statistics revealed that most areas in the country had a lower risk for this outcome compared to the national average. Moreover, the most likely cluster not to have occurred by chance was identified in municipalities in the state of Rio de Janeiro, with a risk 4.7 times higher than the national average, within a radius of 97.8 km (p<0.001) (Figure 1F).

Based on the OLS regression analysis, the Gini Index was positively associated with increased mortality. In contrast, the average number of nurses and the percentage of non-treponemal tests performed on pregnant women were inversely associated with congenital syphilis mortality. The other variables did not remain in the final model. The model’s fit statistics were R2 = 0.006 and AIC = −8,870. The OLS model showed no multicollinearity, with a mean VIF of 2.8. When the same variables were tested in a geographically weighted regression (GWR) model, the direction of the associations remained similar. The GWR model demonstrated improved fit, with R2 = 0.06 and AIC = −8,892, indicating that it was more appropriate for describing the outcome (Table 2).

**Table 2.**
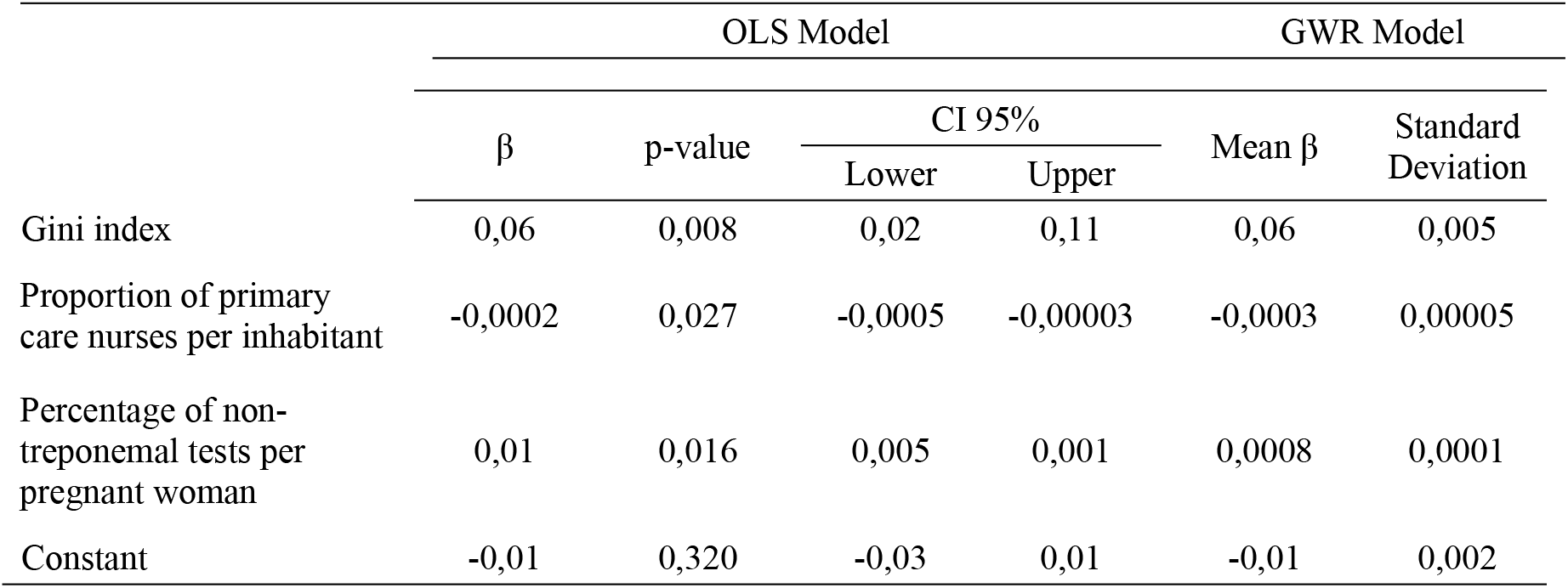
OLS and GWR models for factors associated with congenital syphilis mortality in Brazil, 2008–2022.

| | $\beta$ | p-value | CI 95% | | Mean $\beta$ | Standard Deviation |
| --- | --- | --- | --- | --- | --- | --- |
|  |  |  | Lower | Upper |  |  |
| Gini index | 0,06 | 0,008 | 0,02 | 0,11 | 0,06 | 0,005 |
| Proportion of primary care nurses per inhabitant | -0,0002 | 0,027 | -0,0005 | -0,00003 | -0,0003 | 0,00005 |
| Percentage of non-treponemal tests per pregnant woman | 0,01 | 0,016 | 0,005 | 0,001 | 0,0008 | 0,0001 |
| Constant | -0,01 | 0,320 | -0,03 | 0,01 | -0,01 | 0,002 |

In the geographically weighted model, a positive association was identified between the Gini index and congenital syphilis (CS) mortality, particularly in the states of the Northeast, Southeast, and Central-West regions (Figures 2A and 2B). Conversely, an inverse relationship between the average number of nurses and the outcome was observed throughout the Southeast, South, and Central-West regions (except for Mato Grosso), as well as in the Northeast (except for Piauí and Maranhão) (Figures 2C and 2D). An increase in the availability of non-treponemal tests for pregnant women was associated with a reduction in mortality in the same regions as the previous variable (Figures 2E and 2F).

**Figure 2.**
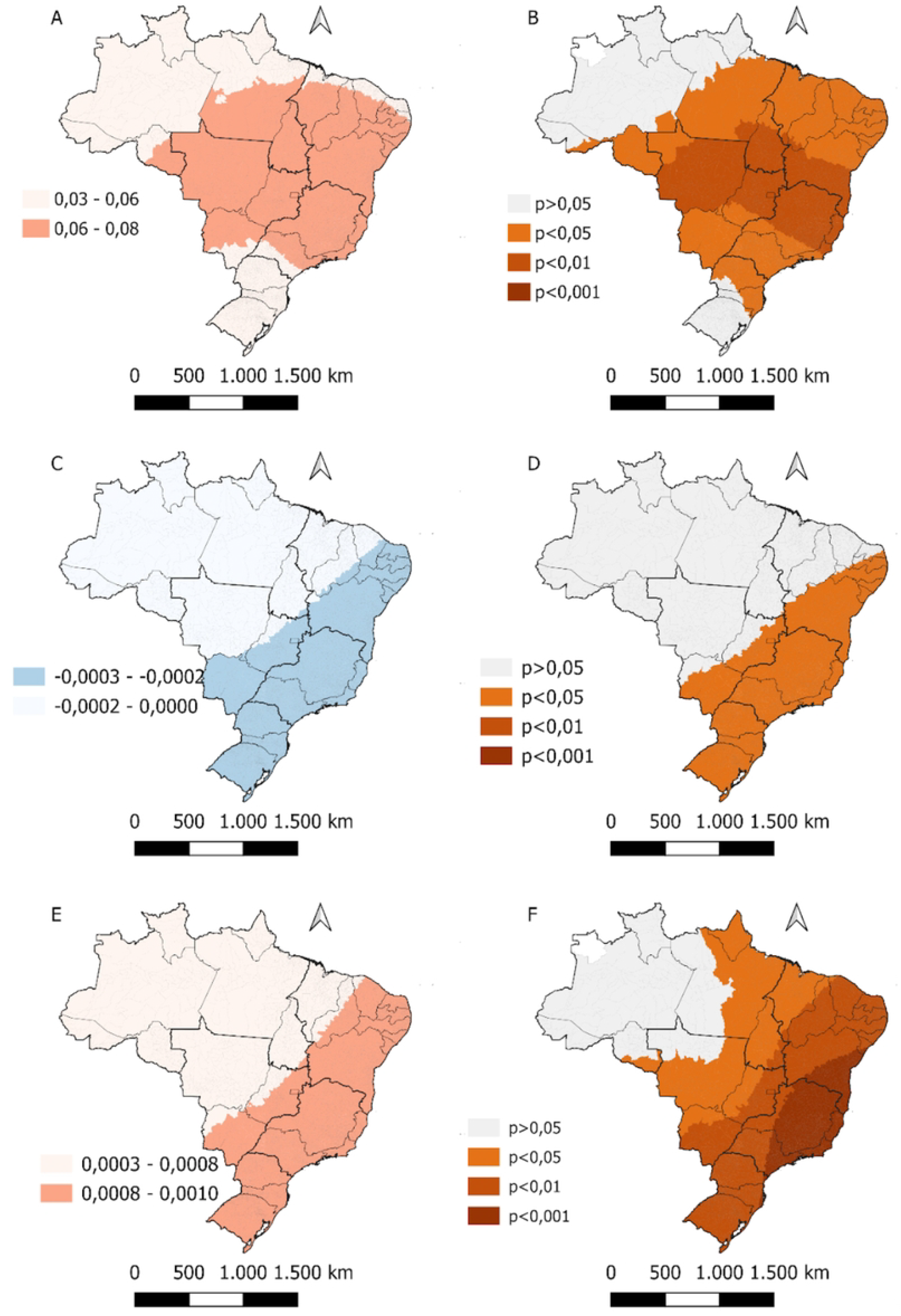
Local indicators associated with congenital syphilis mortality in Brazil from 2008 to 2022.

It is important to highlight that in both models the coefficients were very close to zero and, therefore, should be interpreted with caution. Similarly, the local R2 values were below 10% across the country (Figure 3).

**Figure 3.**
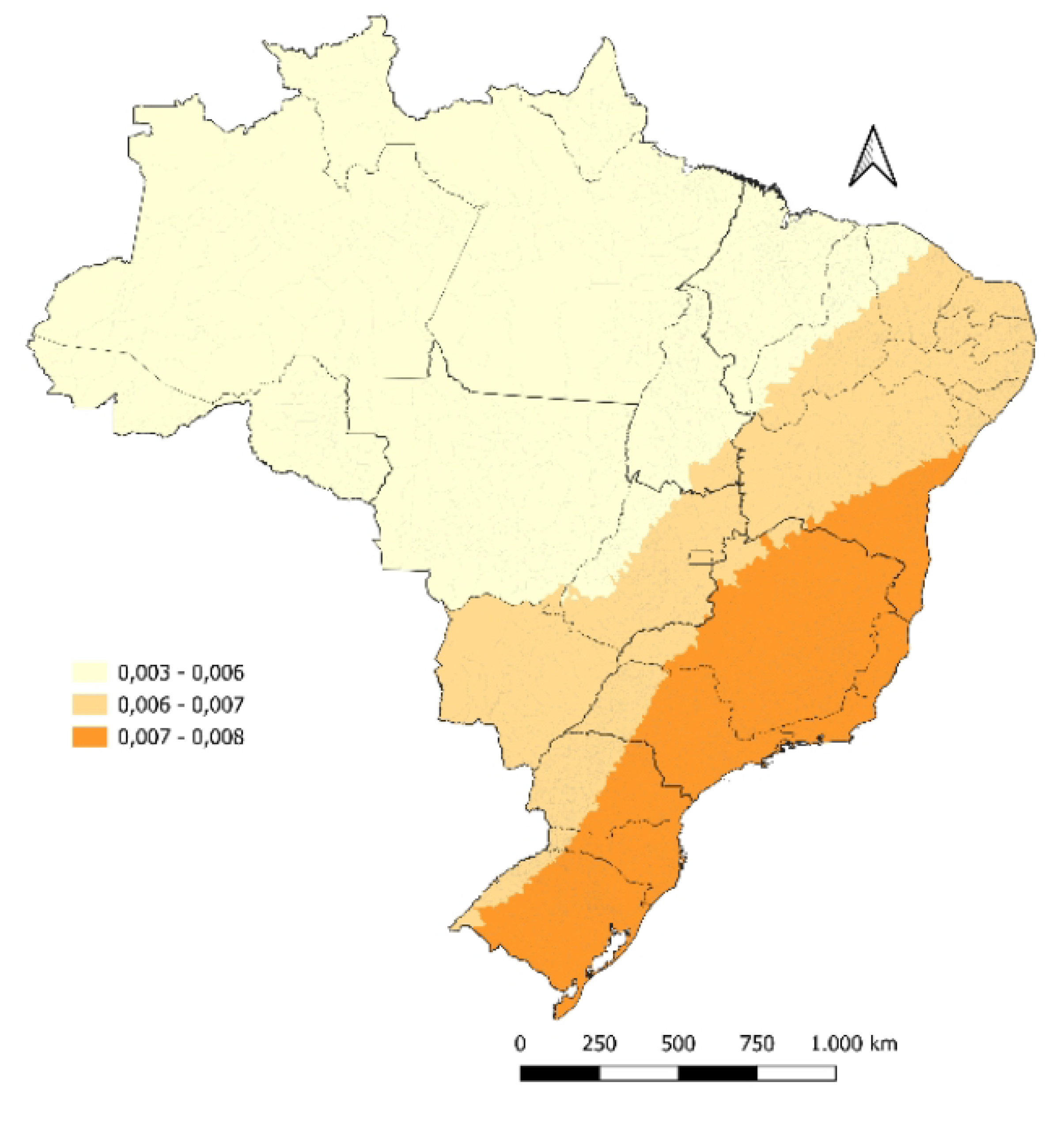
Local coefficient of determination for indicators related to congenital syphilis mortality in Brazil from 2008 to 2022.

## Discussion

Our study analyzed the geographic distribution of deaths due to congenital syphilis (CS) among newborns aged 0 to 27 days in Brazil over a fifteen-year period (2008–2022) and identified associated factors. We observed that most of the country presents a risk of CS-related deaths, particularly in states in the North region (Pará, Acre, Rondônia, and parts of Amazonas) and in Rio de Janeiro. Moreover, mortality is heterogeneously distributed across the Brazilian territory, with the formation of spatial clusters in all states, especially Pará, Rio de Janeiro, and Mato Grosso, which showed a high-high pattern. Finally, we found that the Gini index, the number of nurses in primary care, and the proportion of non-treponemal tests performed on pregnant women are variables that influence the geographic distribution of deaths, particularly in the Northeast, Southeast, Central-West, and South regions.

The Brazilian Ministry of Health reports that both the incidence of CS and related deaths have increased nationwide in recent years [2]. It is estimated that children with CS are twice as likely to die compared to healthy children, which may represent an infant mortality rate (deaths before the age of one year) of 33% [3]. Thus, syphilis remains a significant and ongoing public health issue and one of the main causes of infant mortality, despite being entirely preventable.

In the North region, the largest clusters of CS-related deaths were observed. This is noteworthy given the region’s historical context of underdevelopment and socioeconomic and healthcare inequality [10]. Studies indicate that all states in the North region exhibit poverty indicators above the national average [11], as well as considerable disparities in access to healthcare services (with estimates showing that between 1998 and 2003 all Brazilian regions improved access to healthcare, except the North) [12].

Our findings suggest that the higher probability of CS-related deaths in the North may stem from ineffective control of this condition due to inequities in access to services offered by the public health system (SUS). In this region, barriers to accessing healthcare facilities compromise diagnosis, treatment, and follow-up of syphilis cases, thereby increasing the risk of mortality [4]. Furthermore, the low municipal human development index and limited primary healthcare coverage—especially in Pará and Amazonas—combined with insufficient financial resources, significantly hinder the health care system’s operation. This often results in healthcare services being concentrated in urban areas, limiting access for rural and riverside populations [13].

The state of Rio de Janeiro also showed a high mortality rate due to CS, despite its relatively high level of socioeconomic development. We hypothesize that inadequate treatment of pregnant women or newborns may have contributed to this result. That is, although Rio de Janeiro offers greater access to diagnostic services and better conditions for disease reporting compared to the North region [14], the persistence of inadequate treatment—or lack of adherence by pregnant women—may worsen the condition and increase the risk of death [15,16]. Therefore, controlling CS and related deaths remains a challenge given the multiple influencing factors.

Previous studies have reported a heterogeneous pattern of CS mortality across Brazil, which can be explained by social (income, education, housing, etc.) and healthcare disparities among states, in addition to the country’s vast territorial extension [17,18]. Social conditions— particularly those involving inequities and inequalities—are key risk factors for infant and neonatal mortality, as they affect the allocation of resources for maternal and child healthcare [19]. Thus, a deeper understanding of the problem requires not only an analysis of national public health and social policies, but also state and municipal-level actions, as regional interventions often have greater impact in addressing such conditions [20].

Regarding the inverse association observed between the average number of primary care nurses and the proportion of non-treponemal tests with CS mortality, our findings highlight the need to strengthen primary healthcare. These factors are linked to improved surveillance, and they enable greater preventive and therapeutic assistance for pregnant women and newborns. A systematic review identified the lack of prenatal care as one of the main predictors of neonatal mortality [15]. In addition, it is estimated that appropriate syphilis treatment could reduce neonatal mortality by up to 40% [21]. Therefore, increasing the number of trained nurses in primary care and offering diagnostic testing may reduce barriers to eliminating CS deaths, especially regarding early detection failures and inadequate treatment.

In this study, we also observed that socioeconomic inequality is another key factor predicting CS-related deaths in Brazil. In other words, as inequality—as measured by the Gini index—increases, so do the CS-related deaths. This outcome supports the concept of the social gradient in health proposed by the World Health Organization [22]. Thus, we can affirm that socioeconomic and health conditions vary progressively across territories based on the degree of inequality.

Our findings indicate that CS-related deaths in Brazil demand continued attention from policymakers, health professionals, and researchers. Socioeconomic factors influence mortality both nationally and across macro-regions, suggesting that regional socioeconomic disparities may require targeted actions to control this public health issue [23–25].

The coordination of actions to combat CS at the national level faces several challenges due to Brazil’s continental dimensions [26]. Given the disparities across Brazilian regions, tackling CS requires context-specific public health and social policies, tailored to the socioeconomic and healthcare characteristics of each locality [23].

The information presented in our study helps to identify regions where CS is not being effectively controlled, leading to higher risk of deaths from the disease. On a global scale, these findings may serve as a warning for other countries to enhance surveillance in areas with greater social and health vulnerability, thereby enabling more targeted and timely interventions to prevent newborn deaths from CS.

A limitation of this study lies in the use of secondary data provided by the Brazilian government and the possibility of underreporting. However, we did not observe significant changes in the data collection or reporting structure that would materially affect the results. Finally, we recommend that further studies explore the reasons behind CS-related deaths in regions with better socioeconomic conditions, as these areas would ideally exhibit lower morbidity and mortality due to better access to healthcare services.

## Conclusion

There is a heterogeneous distribution of CS-related deaths in Brazil, characterized by a risk of occurrence in both less developed and more developed regions, as well as the formation of high-high spatial clusters in several states. The distribution dynamics of mortality must be analyzed in light of socioeconomic and healthcare variables, as these are key factors directly influencing mortality patterns across the territory.

## Data Availability

All files are available from the DATASUS database. Link: https://datasus.saude.gov.br/informacoes-de-saude-tabnet/

